# MULTIPLEX PCR FOR SP-D rs3088308 AND CD36 rs1761667 GENE POLYMORPHISMS IN ACTIVE TUBERCULOSIS AND LATENT TUBERCULOSIS PATIENTS

**DOI:** 10.64898/2025.12.03.25341527

**Authors:** Sara Shafqat, Urooj Subhan, Rafia Aslam, Sidra Younis

## Abstract

Tuberculosis (TB) remains a leading cause of morbidity and mortality worldwide, particularly in developing countries such as Pakistan. Despite notable progress in diagnostic strategies, a large proportion of individuals with active tuberculosis (ATB) and latent tuberculosis infection (LTBI) remain undiagnosed, primarily due to limitations in diagnostic accessibility and sensitivity in resource-constrained settings. Genetic variations play a crucial role in determining host susceptibility to TB. Among these, polymorphisms in Surfactant Protein-D (SP-D) rs3088308 and Cluster of Differentiation 36 (CD36) rs1761667 have been associated with differential immune responses in ATB and LTBI. This study was planned to develop and optimize a multiplex Tetra-Primer Amplification Refractory Mutation System PCR (Tetra-ARMS PCR) assay for the simultaneous genotyping of these two polymorphisms in a single reaction; however, the optimization was unsuccessful. We assume the optimization procedure was challenged by factors such as substantial differences in primer annealing temperatures, minimal variation in amplicon sizes, the presence of multiple primers (eight in total), and primer–dimer formation. These constraints limited the assay’s reproducibility and efficiency. To improve multiplexing performance, primer redesign and re-optimization of PCR parameters are recommended. Furthermore, alternative molecular techniques, including high-resolution melting (HRM) analysis or allele-specific PCR, may provide more robust and reliable detection of these polymorphisms within diverse MTB infected populations.

## INTRODUCTIO

TB remains one of the most critical infectious diseases worldwide. It causes major impact on public health and world economy. The disease is caused by a gram-positive bacteria *Mycobacterium Tuberculosis* (MTB). It mainly infects lungs but can disseminate to other organs to cause infection (Boom WH, 2021). According to the WHO, TB is one of the major causes of deaths from infectious diseases across the globe. Incidence of TB is estimated at 134 cases per 100,000 people globally. TB incidence remained disproportionately high in settings with weak healthcare infrastructure, such as Pakistan, one of the top five reporting countries for TB prevalence. Incidence of TB in Pakistan is estimated at 277 cases per 100,000 people, requiring targeted TB control measures (“Global tuberculosis report 2024,” 2024).

ATB is described as a symptomatic infection where the MTB multiply rapidly and leads to clinical symptoms characterized by chronic coughing, weight loss, and also fever. The granuloma when remain in the dormant state in the lungs than the condition is known as the LTBI. Approximately 5-10% of LTBI can convert to ATB due to weak immune system of the host (“Global tuberculosis report 2013,” 2013). The infection begins with the inhalation of MTB-laden droplets that usually reach the alveoli in the lungs. The bacteria attempt to be phagocytized by the resident alveolar macrophages, but the mechanism of microbe destruction is evaded due to the survival and further multiplication of MTB. This results in the formation of granulomas; a structured immune cell unit that contains the infection. A compromised immune response allows the microbes that have been confined in granulomas to break through the mechanism and cause ATB (Scordo, 2016). Additionally, factors like malnutrition or co-infections lead to the reactivation of LTBI.

Genetic factors play a very major role in determining the susceptibility for development of TB. Previous researches show that human genetics involve 25% in development of TB infection. There are many genes involved in development of TB but surfactant protein D (SP-D) gene (Urooj Subhan, 2025) and cluster of differentiation 36 (CD36) gene play a very important role in TB pathogenesis (Ezza Binte Tariq, 2024).

SP-D is a critical component of the innate immune system, playing a vital role in the respiratory defense against pathogens and the maintenance of lung homeostasis (MARIËTTA M. RAVESLOOT-CHÁVEZ1, 2021). In TB, SP-D is involved in recognition of MTB, modulation of immune response and protection against dissemination. It stimulates cytokine production for helping controlling and managing the infection (Sokołowska, 2020). One notable single nucleotide polymorphism (SNP) in the SP-D gene is rs3088308, which has been associated with variations in SP-D expression and function. This polymorphism can affect the protein’s ability to bind pathogens and regulate immune responses, thus influencing susceptibility to lung infections like TB (Hoffmann-Petersen, 2022).

CD36 is member of scavenger receptor family which is involved in TB phagocytosis. The primary expression of CD36 is on macrophages, where it helps in the recognition and uptake of pathogens and regulation of signaling in the immune cells (Wang, 2024). SNP rs1761667 in the CD36 gene have been associated with altered immune responses, potentially impacting the efficiency of bacterial clearance (TARIQ, 2023).

In our lab, studies on genetic variants of SP-D and CD36 genes were performed; where it was confirmed that both of these genes and associated polymorphisms have important role in ATB and LTBI pathogenesis (Ezza Binte Tariq, 2024; Urooj Subhan, 2025).However, performing two different Tetra-ARMS PCR on same sample was time consuming, resource intensive, and laborious. Therefore, we aimed to optimize a multiplex PCR for already designed primers so that variants in both genes could be identified in a single reaction.

## METHODOLOGY

All research was conducted in accordance with the relevant regulations and guidelines approved and reviewed by the Institutional Review Board NUMS (supplementary file S1) according to Declaration of Helsinki. Blood samples from TB patients and TB contacts were collected from the Nishtar Hospital Multan (NHM) and Thalassemia Centre and Safe Blood Bank in Multan, following informed consent. Blood samples for the healthy controls (HCs) were taken from Raza Laboratory Nishtar Road Multan. The sample size for the research was 150 study subjects which included 50 TB contacts, 50 TB patients and 50 HCs. For study subjects, individuals aged 16 or older were included. Patients having HIV infection, those with anemia, and pregnant or nursing mothers were excluded from the study.

The genomic DNA from blood samples (1-2ml) was extracted using the phenol-chloroform method. Already designed and optimized sets of primers for SP-D gene mutation rs3088308 and CD36 gene mutation rs1761667 were used. The details of the primers are given in the supplementary file S2 (Ezza Binte Tariq, 2024; Urooj Subhan, 2025). SP-D gene mutation at rs3088308 and CD36 gene mutation at rs1761667 was amplified using multiplex Tetra-ARMS PCR and PCR products were separated on 2.5% agarose gel.

## RESULTS

### Tetra ARMS PCR for SNP rs3088308 in SP-D

Tetra-ARMS PCR conditions for SNP rs3088308 are given in table 1. Figure 1 shows the representative agarose gel for genotype pattern among controls, patients, and contacts. In the homozygous wild type TT genotype, there were two bands (515 bp+347 bp). The heterozygous AT genotype has three bands: 515 bp, 347 bp, and 217 bp. On the other hand, the homozygous mutant genotype AA showed two bands, each of 515 bp and 217 bp length.

**Table 1.**
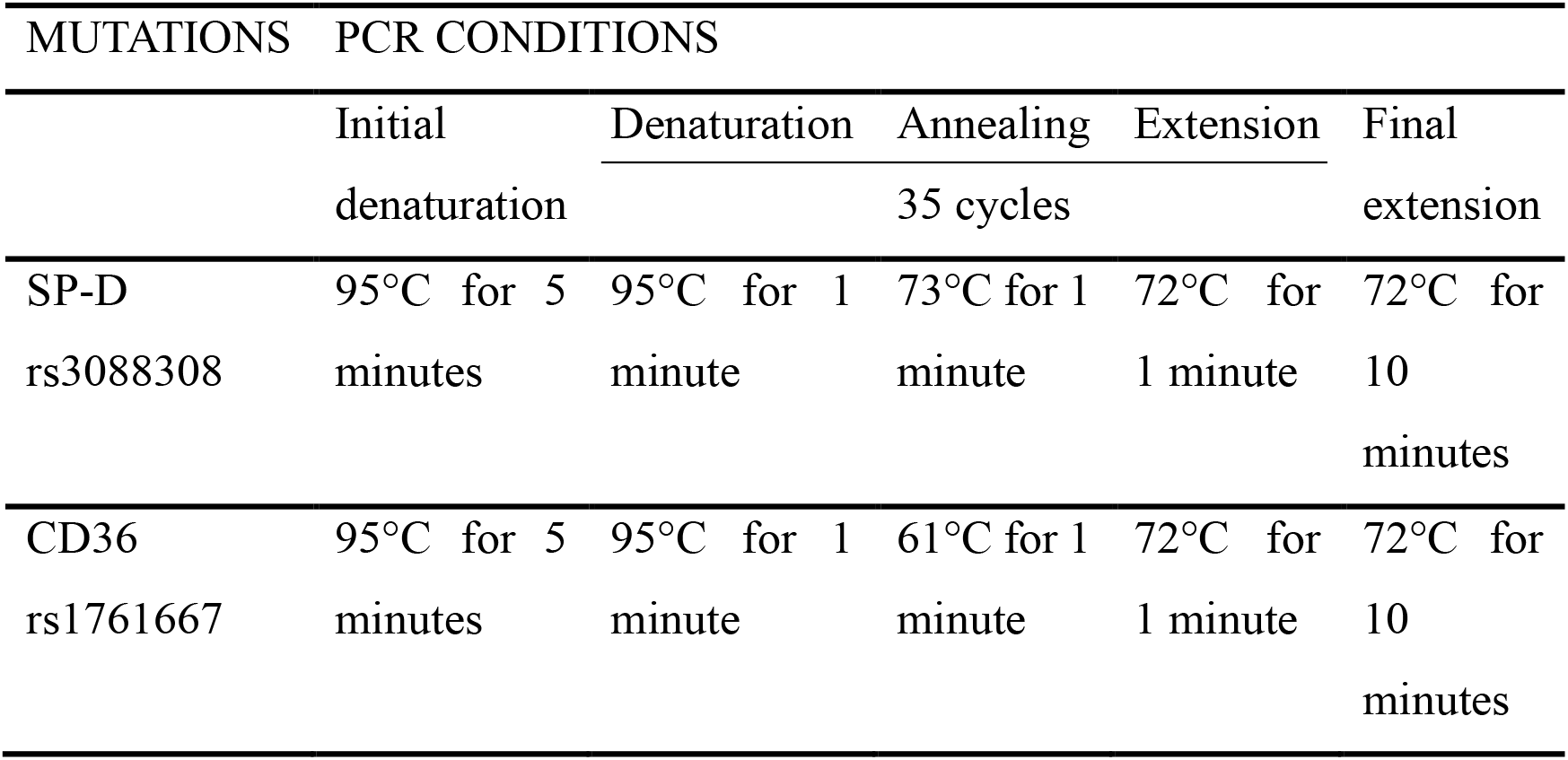
Tetra-ARMS PCR conditions for SP-D gene mutation rs3088308 and CD36 gene mutation rs1761667.

**Figure 1.**
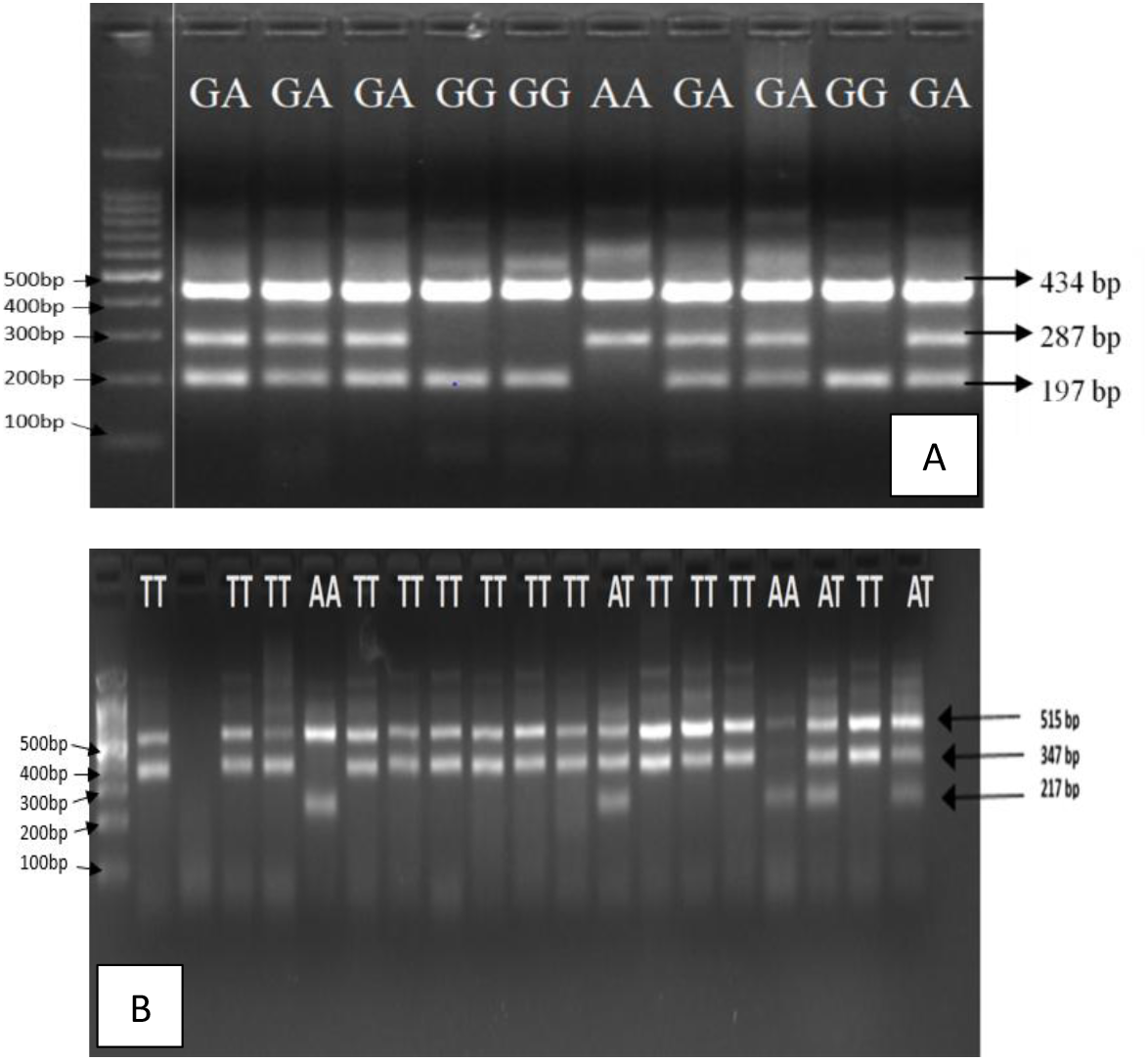
Agarose gel picture representing genotype pattern for SNP rs1761667 (A) and SNP rs3088308 (B)

### Tetra ARMS PCR for SNP rs1761667 in CD36

Tetra-ARMS PCR conditions for SNP rs1761667 are given in table 1. Figure 1 shows the representative agarose gel for genotype pattern among controls, patients, and contacts. In the homozygous wild type GG genotype, there were two bands (434 bp+197 bp). The heterozygous GA genotype has three bands: 434 bp, 287 bp, and 197 bp. On the other hand, the homozygous mutant genotype AA showed two bands, each of 434 bp and 287 bp length.

### Multiplex PCR optimization for the SNP rs3088308 and rs1761667

Conditions for multiplex PCR for the SNP rs3088308 and rs1761667 are given in table 2. Figure 2 shows the representative agarose gel for genotype pattern for multiplex PCR. There were no visible DNA bands on temperatures 65.5°C, 70.7°C, and 72.1°C. On temperature 61.9°C, there were three visible bands of 434bp, 347bp, and 197bp. Two bands 434bp and 197bp indicated genotype pattern for SNP rs1761667 and there were only one band 347bp for SNP rs3088308.

**Table 2.**
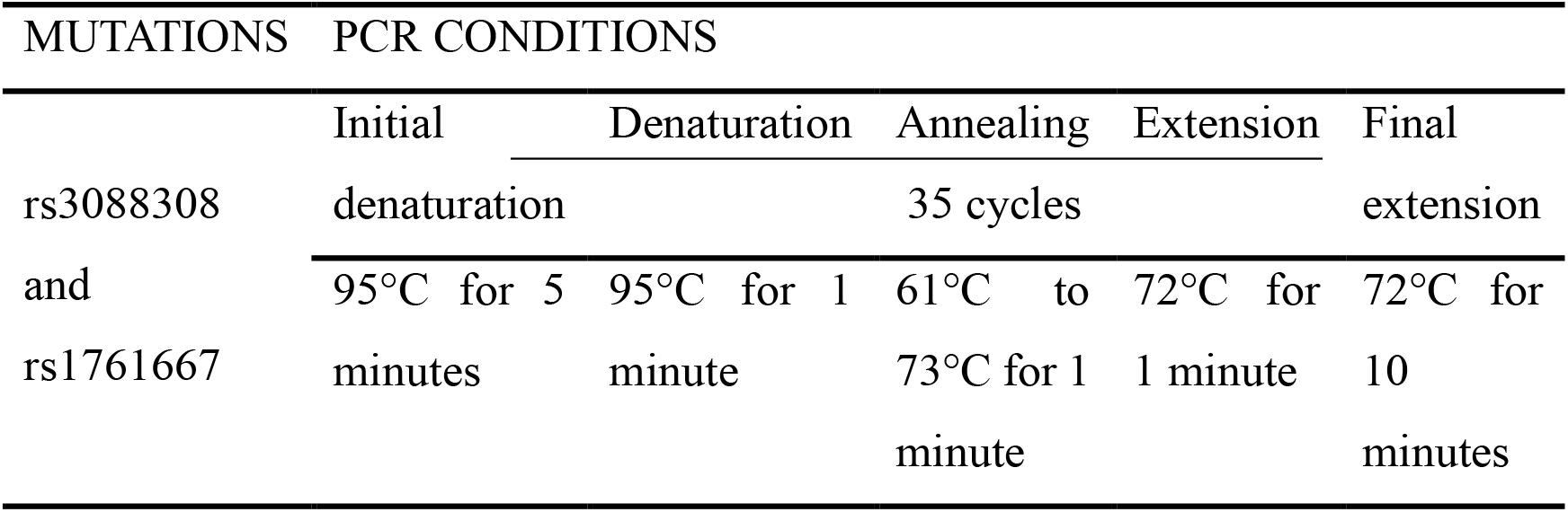
Multiplex PCR conditions for SP-D gene mutation rs3088308 and CD36 gene mutation rs1761667.

**Figure 2.**
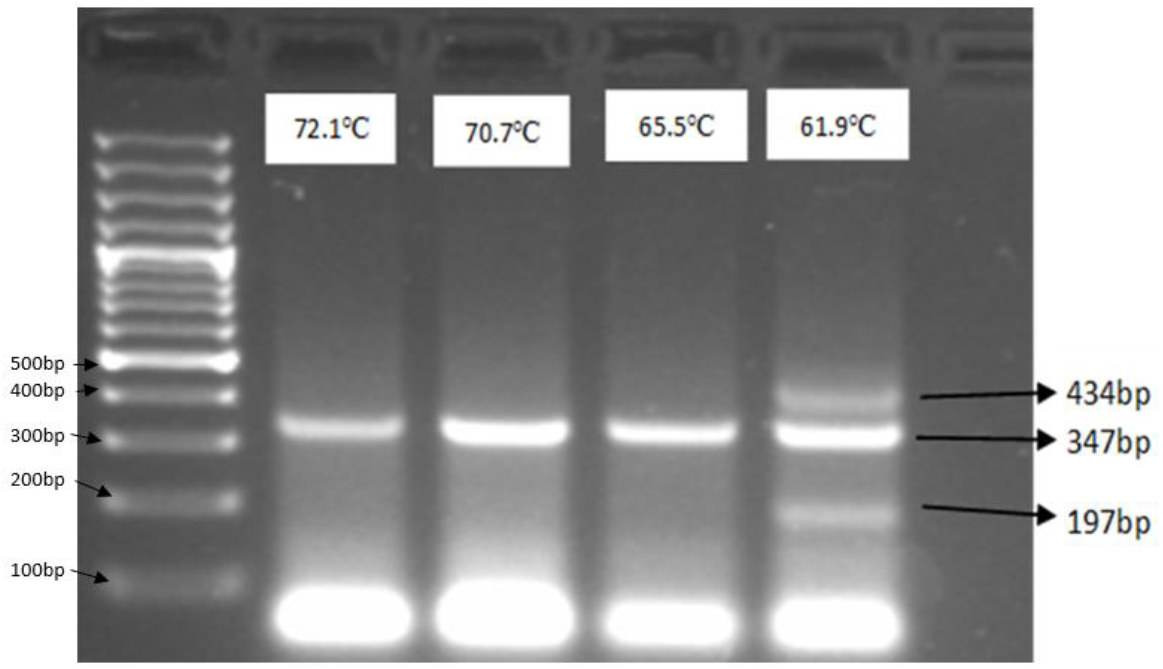
Agarose gel picture representing genotype pattern for multiplex PCR

## DISCUSSION

We aimed to optimize multiplex Tetra-ARMS PCR conditions for SNPs, SP-D rs3088308 and CD36 rs1761667. But there were several challenges for optimization of multiplex PCR for CD36 rs1761667 and SP-D rs3088308. Precise temperature control is crucial for the degree of amplification in PCR that even a slight variations, like 0.5°C, might affect the banding patterns seen in gel electrophoresis (YANG, 2005). The annealing temperatures of the primers for the two SNPs were 61°C (rs3088308) and 73°C (rs1761667) and had significant difference of 12°C, which made the amplification on same temperature very challenging. Furthermore, when multiple primers in single PCR amplify different targets simultaneously, issues such as primer dimers or inefficient primer binding could result in some regions being overrepresented or underrepresented. In single reaction mixture of multiplex PCR, the total number of primers was 8 and it could result in formation of primer dimers in reaction which may lead to coverage bias (ITOKAWA, 2020). Additionally, resolution of PCR products is a challenge especially when separating products of smaller and comparable sizes. 2.5% agarose gel might not have had enough resolving ability to separate bands with smallest size differences, even though it offered somewhat better resolution for smaller DNA fragments.

We recommend purposed based re-designing of the primer sets for these SNPs and carefully considering annealing temperature difference can be done to avoid primer dimerization and improve amplification of sample DNA. Agarose gel can be substituted with acrylamide gel, for even distinguishing between DNA fragments that differ by just one base pair. Alternative molecular techniques, including high-resolution melting (HRM) analysis or allele-specific PCR, can be considered for reliable detection of these polymorphisms.

Our study had some limitations. We had only focused on optimization of temperature in multiplex Tetra-ARMS PCR, while other conditions like chemical composition of mixture, primer redesigning, and PCR cycle time was not changed. Other modifications of the PCR, for example Touchdown PCR, Hot start PCR, and High-resolution melting PCR were not considered.

## Data Availability

All data produced in the present work are contained in the manuscript.

## ACKNOWLEDGEMENTS

We acknowledge the study subjects who have voluntarily provided their blood samples for this investigation.

## AUTHORS’ CONTRIBUTIONS

Sara Shafqat: Data Curation, Investigation, Writing Original Draft

Urooj Subhan: Writing-Review and Editing

Rafia Aslam: Investigation

Dr. Sidra Younis: Conceptualization, Project Administration, Methodology, Supervision, Writing-Review and Editing

