## Supplementary file for "MULTIPLEX PCR FOR SP-D rs3088308 AND CD36 rs1761667 GENE POLYMORPHISMS IN ACTIVE TUBERCULOSIS AND LATENT TUBERCULOSIS PATIENTS"

#### S1. Approval certificates and Questionnaires

**DEPARTMENT OF BIOCHEMISTRY AND BIOTECHNOLOGY**
**THE WOMEN UNIVERSITY, MULTAN**

Date: 03-03-2023

To,
Medical Superintendent
Nishtar Hospital Multan.

Subject: <u>Sampling for a research project titled 'Scavenger a receptors genes polymorphisms association with active and latent tuberculosis in Pakistani population'</u>

1. The subject project has been approved by NUMs IRB for Dr. Sidra Younis research. Ms. Urooj Subhan and Ayesha Yousaf are working as a researcher on this project under the supervision of undersigned and co-supervision of Dr. Sidra Younis. To commence the project research work, they need to collect 3-5ml blood samples from TB patients (n=100) and TB contacts (n=100). The sampling duration will be 2 months.
2. Keeping above in the view, it is requested that MPhil scholars, Ms. Urooj Subhan and Ms. Ayesha Yousaf may please be allowed to collect blood samples for the said project.

Kindly grant permission to do so.

Dr. Farah Deeba
Assistant Professor
Department of Biochemistry & Biotechnology
The Women University Multan

I have no objection if Hospital/NMU administration allows the said Research.

Prof. Dr. M. Azam Mushtaq

F/w to [illegible] Admin Registrar [illegible]

Medical Superintendent
NISHTAR HOSPITAL MULTAN

7/3/23

NUMS
NATIONAL UNIVERSITY OF MEDICAL SCIENCES

National University of Medical Sciences
The Mall Rawalpindi
No: 06 / IRB&EC / NUMS / 24

18 Oct 2022

**CERTIFICATE OF APPROVAL - NUMS-IRB & ETHICAL COMMITTEE**

A research project titled "Scavenger Receptors Genes Association with Active and Latent Tuberculosis in Pakistani Population" in respect of PI-Dr Sidra Younis, Assistant Professor, Department of Biological Sciences, NUMS is hereby considered to be exempted for review by the IRB & Ethical Committee, NUMS.

Assistant Director ORIC
Secretary IRB & Ethical Committee
**(Aqeel Yunus)**

**COUNTERSIGNED**

Maj Gen
Pro-Vice Chancellor (Acad)
Chairman, NUMS-IRB & Ethical Committee
**{Saleem Ahmed Khan, HI(M) (Retd)}**

To: NDBS (For PI – Dr Sidra Younis)

### TB genetics Pakistan CRF: Active TB patients

#### 1. PERSONAL DETAILS

First name

Last name

Address

Mobile number

**Alternative contact**:

FAMILY MEMBER / FRIEND THAT MAY BE CONTACTED

Contact person/relationship /

Contact person mobile number

#### 2. STUDY VISIT DETAILS AND ELIGIBILITY

Date of visit ______________________________ ____________________

**Eligibility:**

| **Inclusion criteria** | **Yes** | **No** |
|---|---|---|
| 1. Aged 16 or above | | |
| 2. Newly diagnosed active TB about to start treatment | | |
| 3. Gives written informed consent to participate in the study | | |
| **Exclusion criteria** | | |
| 1. Known Anaemia (Hb <10 g/dl) | | |
| 2. Known HIV infection | | |
| 3. Pregnant or breastfeeding | | |

**Note**:
* If the answer to any of the inclusion criteria is **NO**, the subject is not eligible for the study *
**If the answer to any of the exclusion criteria is **YES**, the subject is not eligible for the study**

#### 3. DEMOGRAPHIC AND CLINICAL DETAILS

| | |
|---|---|
| Age | |
| Gender | |
| Ethnic origin | |
| Site of disease | |
| Basis of diagnosis (Smear, GeneXpert, other) | |
| Duration of symptoms | |
| Organism isolated | |
| Drug-susceptibility | |
| Previous anti-TB treatment | |
| Details if so | |
| Past medical history | |
| Current medications | |
| Smoking history | ○ Current ○ex-smoker ○never |
| BCG status (BCG scar present or absent?) | |

#### 4. RESULTS

| **Laboratory test done** | **Result** |
|---|---|
| Haemoglobin level if available [state N/A if not available] | |
| Chest x-ray if available [state N/A if not available]<br>Date: | |

**Actual volumes (mL) of blood collected:**

| **Tube** | **Target volume** | **Actual volume** |
|---|---|---|
| EDTA | 5 mL | |

Date and time of blood collection: ______________________________________________

Date and time of receipt of blood in laboratory: __________________________________

Received by (name of laboratory personnel): _____________________________________

**Details of person completing CRF:**

Name__________________________Signature ____________________________________

### TB genetics Pakistan CRF: Healthy controls

#### 1. PERSONAL DETAILS

First name

Last name

Address

Mobile number

**Alternative contact:**

FAMILY MEMBER / FRIEND THAT MAY BE CONTACTED

Contact person/relationship /

Contact person mobile number

#### 2. STUDY VISIT DETAILS AND ELIGIBILITY

Date of visit ____________________ ____________

**Eligibility:**

| **Inclusion criteria** | **Yes** | **No** |
|---|---|---|
| 1. Aged 16 or above | | |
| 2. Doesn't have symptoms of TB | | |
| 3. Gives written informed consent to participate in the study | | |
| **Exclusion criteria** | | |
| 1. Is diagnosed with TB/ has been in contact with TB patient | | |
| 2. Known Anaemia (Hb <10 g/dl) | | |
| 3. Known HIV infection | | |
| 4. Pregnant or breastfeeding | | |

**Note**:

* If the answer to any of the inclusion criteria is **NO**, the subject is not eligible for the study *

**If the answer to any of the exclusion criteria is **YES**, the subject is not eligible for the study**

**3. DEMOGRAPHIC AND CLINICAL DETAILS**

| | |
|---|---|
| Age | |
| Gender | |
| Ethnic origin | |
| Previous anti-TB treatment | |
| Details if so | |
| Past medical history | |
| Current medications | |
| Smoking history | ○Current ○ex-smoker ○never |
| BCG status (BCG scar present or absent?) | |

**4. RESULTS**

| Laboratory test done | Result |
|---|---|
| Haemoglobin level if available [state N/A if not available] | |
| Chest x-ray if available [state N/A if not available]<br>Date: | |

**Actual volumes (mL) of blood collected:**

| Tube | Target volume | Actual volume |
|---|---|---|
| EDTA | 5 mL | |

Date and time of blood collection: ______________________________________________

Date and time of receipt of blood in laboratory: ___________________________________

Received by (name of laboratory personnel): _______________________________________

**Details of person completing CRF:**

Name__________________________Signature ____________________________________

### TB genetics Pakistan CRF: TB Contacts

#### 1. PERSONAL DETAILS

First name [ ]

Last name [ ]

Address [ ]

Mobile number [ ]

Alternative contact:

FAMILY MEMBER / FRIEND THAT MAY BE CONTACTED

Contact person/relationship [ / ]

Contact person mobile number [ ]

#### 2. STUDY VISIT DETAILS AND ELIGIBILITY

Date of visit ______________________

**Eligibility:**

| **Inclusion criteria** | **Yes** | **No** |
|---|---|---|
| 1. Aged 16 or above | | |
| 2. Household contact with index case of pulmonary TB within the last 6 months | | |
| 3. Asymptomatic (*i.e.,* no cough, weight loss, fever, night sweats, lymphadenopathy or any other symptom / sign of active TB) | | |
| 4. Gives written informed consent to participate in the study | | |
| **Exclusion criteria** | | |
| 1. Clinical suspicion of active TB | | |
| 2. Known anaemia (Hb <10 g/dl) | | |
| 3. Pregnant or breastfeeding | | |
| 4. Known HIV infection | | |

**Note**:

* If the answer to any of the inclusion criteria is **NO**, the subject is not eligible for the study *

**If the answer to any of the exclusion criteria is **YES**, the subject is not eligible for the study**

**3. DEMOGRAPHIC AND CLINICAL DETAILS**

| | |
|---|---|
| Age | |
| Gender | |
| Ethnic origin | |
| Occupation | |
| Place of contact with index pulmonary TB case | ○ Household<br>○ Other |
| Index case's relationship with the participant | ○ Parent/sibling<br>○ Other |
| Participant's proximity to index case | ○ Sleep in the same bed<br>○ Sleep in different bed but same room<br>○ Sleeps in a different room, same house<br>○ Sleeps in a different house |
| Activities shared with the index case | ○ Every day, >=50% of the day<br>○ Every day, <50% of the day<br>○ Not every day |
| How long was the index case coughing before their anti-TB treatment was started? | |
| Past medical history | |
| Current medications | |
| Smoking history | ○ Current ○ ex-smoker ○ never |
| BCG status (BCG scar present or absent?) | |

**4. RESULTS:**

| Test done | Result |
|---|---|
| Haemoglobin level if available [state N/A if not available] | |

**Actual volumes (mL) of blood collected:**

| Tube | Target volume | Actual volume |
|---|---|---|
| EDTA | 5 mL | |

Date and time of blood collection: ________________________________________________ Date and time of receipt of blood in laboratory: ________________________________________

Received by (name of laboratory personnel): ______________________________________

**Details of person completing CRF:**

Name__________________________Signature ___________________________________

**S2. Sequence and properties of primers of SP-D rs3088308 and CD36 rs1761667**

| Gene | Mutation | Primers | Sequence | Length (bps) | Tm (℃) |
|---|---|---|---|---|---|
| SP-D | rs3088308 | Outer forward | GTGGGGAATCATGGT GTCACCTCACAGGGT | 30 | 73 |
| | | Outer reverse | CCACACAGTCCTCTG ACCCGCCATCATC | 28 | 72.2 |
| | | Inner forward | CACACAGGCTGGTGG ACAGTTGGGCA | 26 | 72.7 |
| | | Inner reverse | GCATTCTCAGCGGCA GAGCGTGGTGA | 26 | 70.9 |
| CD36 | rs1761667 | Outer forward | AAGGTCTGGTATCCAC CTGTTTTCCT | 26 | 64.7 |
| | | Outer reverse | AAGAGTTTTCATGAA GCTTCCCGC | 24 | 61.9 |
| | | Inner forward | TTTTATTCATCTTTGCA TGCCATCG | 25 | 57.3 |
| | | Inner reverse | TCATACTCCAGGCTTT GAGCATTGT | 25 | 61.9 |
